# Quantifying and Visualizing Emergency Physician Workflow: Results of an Observational Time-Motion Study

**DOI:** 10.1101/2024.11.27.24318109

**Authors:** Andrew J. Henreid, Kimon L.H. Ioannides, Joshua M. Pevnick, Tara N. Cohen, Sam S. Torbati, Teryl K. Nuckols, Carl T. Berdahl

## Abstract

**Background:** Emergency physicians face considerable workflow challenges in the emergency department (ED) due to unpredictable work environments, frequent interruptions, and mounting documentation requirements. Excessive time away from direct patient care is increasingly viewed as detrimental to care quality, communication, and patient safety.

**Objective:** This study aimed to quantify and visualize how emergency physicians allocate their time during ED shifts, particularly time spent on the computer.

**Methods:** This observational time-motion pilot study was conducted in a high-volume, urban ED. We used a specialized application to continuously track physician activities including computer use, direct patient care, communication, and all other tasks carried out on shift. Electronic health record (EHR) event log data were queried to estimate physicians’ computer use after their scheduled shift. The main outcome was the proportion of time spent on each activity. The primary measure was total minutes of computer use (during and after shift) per scheduled hour of clinical work.

**Results:** We observed 20 emergency physicians for one 8–9 hour clinical shift each, generating over 150.0 hours of real-time observation data quantifying physicians’ ED workflow. Physicians spent a median 34.1% of their shift using the computer and 26.9% with patients. Other activities included verbal communication with staff (15.9%), phone use (9.5%), miscellaneous tasks (5.5%), personal time (3.9%), electrocardiogram review (0.7%), and procedures (0.4%). EHR log analysis showed that physicians spent an additional median 1.3h (IQR 0.5–2.6) using the computer after their scheduled shifts, for a combined 29.8 minutes (IQR 25.6–38.5) on the computer per scheduled hour of their ED shift.

**Conclusions:** Emergency physicians spent more than one-third of their ED shift working on the computer, which was more time than they spent with patients. They also spent 1–2 hours using the computer after their scheduled shift. These findings demonstrate the need for strategies to reduce unnecessary computer use during and after clinical shifts to enhance workflow efficiency and improve patient care.

## Introduction

### Background

There is rising concern that too much time spent away from the bedside may undermine quality of medical care due to decreased communication and fewer opportunities to directly interact with patients [1-3]. Recent studies of physician workflow in the United States and internationally have concluded that frequent interruptions and burdensome documentation requirements place patients at risk for harm [4-6]. The emergency department (ED) setting is a particularly challenging work environment for physicians given its inherent unpredictability [7]. Prior literature has demonstrated that interruptions in the ED are persistent, and that task switching can jeopardize patient safety [8-10].

The application of methods from human factors engineering, such as time-motion studies, can help identify inefficiencies, optimize performance, and improve work design [11-13]. In preparation for tests exploring the impact of tools designed to reduce time at the computer in the ED, we sought to establish baseline measurements.

### Objectives

The objective of this pilot study was to assess and characterize emergency physician workflow by quantifying the time ED physicians allocate to clinical and non-clinical tasks during their shifts. The main outcome was the proportion of time spent across each type of observable activity, with the primary measure being total minutes of computer use per scheduled hour of the physician’s shift. Secondary outcomes included the percentage of time spent on specific activity categories, including computer use, interacting with patients, verbal communication with clinical staff, operating the phone, reviewing electrocardiograms (EKG), performing medical procedures, personal time, and all other nonspecific miscellaneous tasks.

## Methods

### Study Design & Setting

We conducted an observational time-motion study and review of electronic health record (EHR) event logs to track emergency physician activities in an urban, high-volume ED that is a Comprehensive Stroke Center, ST Elevation Myocardial Infarction-Receiving Center, and Level-1 Trauma Center with an annual census of 90,000 ED visits. Observations were conducted between May 21 and November 6, 2019. During the study period, attending emergency physicians supervised or provided care for all patients. Residents from non-emergency medicine specialties (e.g., internal medicine, anesthesia, etc.) assisted with care for less than 5% of all visits, and physician assistants were involved with 20%. The number of emergency physicians staffing the ED varied from a minimum of 3 during overnight shifts to a maximum of 6 during peak hours.

### Recruitment

Physicians were recruited through convenience sampling and consented to the study in a private setting prior to their observed shift. Participant eligibility criteria included emergency physicians credentialed to practice in the institution’s ED. No other exclusion criteria were used for this study sample.

### Ethical Consideration

This study involving human participants was approved by the Institutional Review Board of Cedars-Sinai Medical Center (Pro00057066) prior to data collection. Written informed consent was obtained from each participating physician before their observed shift [14]. The institutional review board approved a waiver of consent for patients, as the primary focus of the study was to observe physician activity where direct contact with patients and sensitive information was expected to occur only incidentally. All collected data were stored on secure institutional servers. Participants did not receive compensation for this study.

### Measures

The main outcome was the proportion of scheduled shift time spent in each activity category. The primary measure of interest was the total minutes of computer time per scheduled hour of the physician’s clinical shift, defined as the sum of observed on-shift computer use and after-shift computer use from EHR event logs divided by scheduled shift hours. Calculation of total computer time included the number of minutes each physician spent on the computer, both during and immediately after their scheduled ED shift. Secondary outcomes measures included the percentage of time spent on specific shift activities, including computer usage (Computer), direct patient interaction (Patient), verbal communication with clinical staff (Verbal), operating the phone (Phone), reviewing EKGs (EKG), and performing procedures (Procedures).

EHR event log data provided detailed timestamps for all user actions carried out within the EHR computer system (e.g., “ED Disposition Activity viewed,” “User logged in,” “Notes viewed,” etc.) after each observed shift. *EHR use* was defined as the number of seconds each physician spent *actively* using the EHR during their event log timespan, using a 1-minute threshold of inactivity [15]. Physician EHR use was thus considered inactive if more than 59 seconds elapsed between any two successive EHR actions, indicating the physician left the workstation or diverted their attention to perform other tasks [15, 16]. *Total active use* was determined by calculating the time intervals between access times (access instants) for every active EHR action (i.e. the number of seconds between every two subsequent logged events).

*Pajama Time* is a term attributed in academic literature to characterize physician time spent using the EHR (computer) after the end of their scheduled shift (i.e., on administrative actions such as writing clinical notes) [17-19]. In this study, we defined pajama time as any EHR use that occurred within two days after in-person observation concluded or before the physician’s next clinical shift [17, 18, 20]. Pajama time was measured using EHR event logs capturing EHR actions up to 48 hours after the observed shift day (two-day span), which included EHR use occurring either on-site at the medical center or elsewhere (i.e., at home). *See Figure 1. Overview of Data Collection Process* for a depiction of the combined data collection methods utilized in this study.

**Figure 1.**
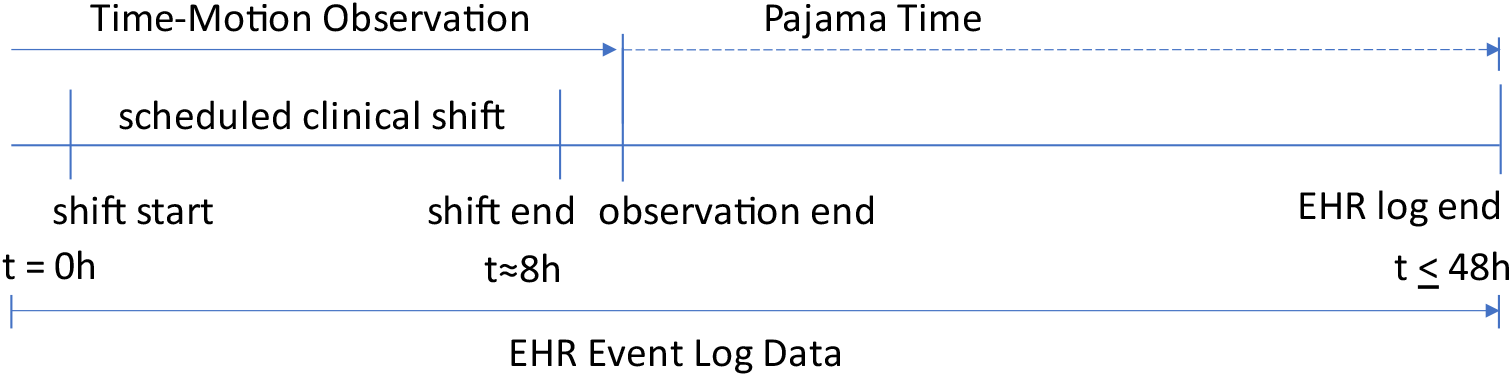
Overview of Data Collection Process. Figure depicts the data collection timeline beginning with in-person observation (Time-Motion Observation) at the start of each physician’s shift (t = 0h), followed by pajama time (t = 8h–48h) captured through EHR Event Log Data. *\*Pajama time defined as EHR use after the observation period ended, where t represents relevant time markers in the data collection period (ranging from 0 hour when the physician began their shift to 48 hours proceeding their observed shift)*.

### Data Collection

#### Time-motion observation data

In-person, time-motion observations were carried out to collect workflow data characterizing ED physician time spent on pre-specified tasks during ED shifts (one observed shift per enrolled physician). A single, trained observer with experience in clinical care as an emergency medical technician (AJH) shadowed enrolled emergency physicians during clinical shifts that were scheduled to last 8 or 9 hours. The observer began data collection at the time the shift began and ended only when the physician informed the observer that they were done interacting with patients and planned to either leave the ED or use the computer without further patient interaction. The observer took a 45-minute break from data collection approximately halfway into each shift and length of shift was adjusted to account for this period prior to analysis.

The trained observer used a digital tablet and web-based, time-motion application (*TimeCaT 3*.*9*) to track start and end times (to the nearest second) across a list of pre-specified tasks [21]. Time-motion studies are a well-established methodology for workflow research and are considered “gold standard methods” for healthcare observations [11]. Similarly, the TimeCaT application used for in-person data collection was specifically designed and validated for measuring workflow in healthcare contexts [21]. *See Figure 2. TimeCaT Data Capture Interface* for the graphical user interface used for in-person observation data collection.

**Figure 2.**
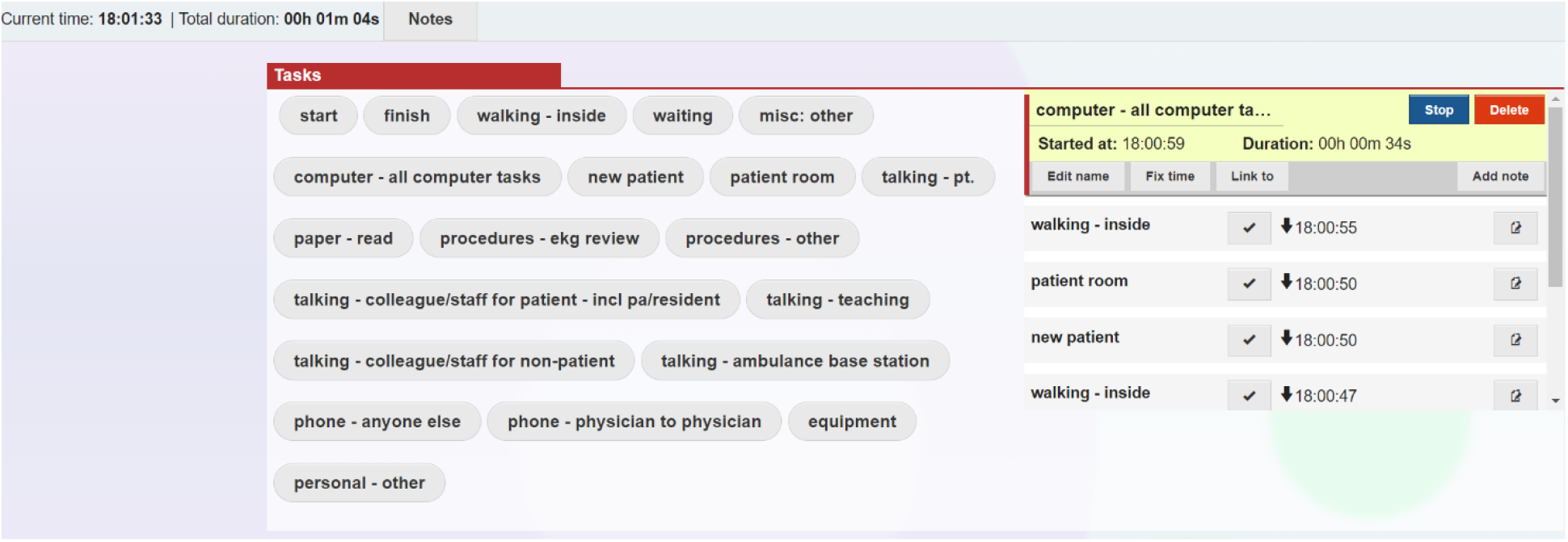
TimeCaT Data Capture Interface (Time-Motion) Figure presents an example of the TimeCaT 3.9 interface used for in-person observation data collection (time-motion)from the observer’s perspective

Physician workflow (defined as the series of activities carried out by the physician during their shift) [22] was continuously assigned to one dominant task by clicking an activity-specific button in the TimeCaT application. These tasks included computer use, direct patient interaction, variations of phone use, EKG review, other clinical procedures, verbal communication with hospital staff, verbal instruction to staff, reading or writing on paper, waiting for next task, gathering equipment, walking within the medical center, personal time, and miscellaneous actions. For clarity and comprehension, individual tasks were then combined into the following activity categories: computer use (Computer), patient interaction (Patient), verbal communication with ED staff (Verbal), operating the phone (Phone), EKG review (EKG), clinical procedures (Procedures), all personal tasks (Personal), and any remaining miscellaneous tasks (Miscellaneous). ***See*** *Table 1. Categorization of Physician Activities* for details on how individual tasks were classified into major activities.

**Table 1.** Categorization of Physician Activities (Time-Motion)

| ACTIVITY CATEGORY | INDIVIDUAL TASK |
| --- | --- |
| Computer | computer - all computer tasks |
| EKG | procedures - EKG review |
| Misc | misc: other / other<br>paper – read<br>Waiting<br>walking – inside |
| Patient | new patient (*unused*)<br>patient room<br>talking - pt. |
| Personal | Equipment<br>personal – other |
| Phone | phone - physician to physician<br>phone - anyone else |
| Procedures | procedures - other (*i.e., non-EKG*) |
| Verbal | talking - colleague/staff for patient - incl pa/resident<br>talking – teaching<br>talking - colleague/staff for non-patient<br>talking - ambulance base station |
| *Excluded research admin tasks* | Start<br>Finish |

When physicians were performing multiple tasks simultaneously (e.g., talking on the phone while using the computer), the observer determined which task was the central focus of the physician’s attention and alternated accordingly. This, in addition to the highly variable nature of ED workflow, resulted in many transitions between tasks during periods of multitasking, which the TimeCaT software was designed to capture [21]. When the physician entered patient rooms, the observer waited outside the room in the interest of patient privacy, and time spent inside the patient’s room was assumed to be spent on direct patient care.

#### Event log data from the electronic health record

To supplement the time-motion data collected during in-person observations, we also obtained EHR event log data to estimate the amount of time physicians spent on the EHR after the end of each physician’s observed shift [16, 23, 24]. Each event log contained a detailed list of actions performed within the EHR by a particular user over a specified period of time, and each action is accompanied by a time-stamp [23-27]. We collected the EHR event log data from the time in-person observation ended until up to 48 hours later or the time that the physician’s next shift began (whichever was earlier). One physician’s EHR event log data was excluded from “pajama time” analysis because their unique administrative responsibilities suggested that their EHR actions were not directly related to their clinical care.

### Analysis

Descriptive analyses were performed using Microsoft Excel and SPSS Statistics (Version 24.0). Data visualizations were created using Tableau Desktop, Microsoft Office, and Seaborn [28]. Data visualizations were used to illustrate physician workflow allocation by activity type and sequence of activities over the course of each ED shift.

## Results

### Overview

Twenty attending emergency physicians were enrolled in the study (5 female, 15 male). The median number of years in practice since residency graduation was 14.5 (IQR 7.5– 23.0; range 3 to 44 years), with a range in residency graduation year of 1976 to 2017. We collected a total of 150.0 hours of time-motion data, with a median observation duration of 7.3h (IQR 7.2–8.1) per physician. This included a total of 14 eight-hour shifts and 6 nine-hour shifts with a shift start time range of 5:00 AM–1:00 PM.

EHR event log data were obtained for all 20 clinical shifts, starting from the time the shift observation ended and continuing for up to 48 hours or until the beginning of the physician’s next clinical shift. Initially, our dataset of EHR event logs included descriptions of physician actions over a total of 200.3h including pajama time. After exclusion of one physician with unique administrative responsibilities, EHR event log data used in the reported analysis included a total of 189.1h [median 10.1h (IQR 8.5– 10.6) per physician] across the remaining 19 physicians in the sample.

### Outcomes

For the primary outcome measure of total computer time per scheduled hour of clinical shift, we found that ED physicians spent a median 29.8 minutes (IQR 25.6–38.5) on the computer per each hour of scheduled clinical work. Total minutes spent on the computer included both directly observed computer time during the ED shift and EHR pajama time that occurred after the physician’s observed shift.

#### Direct Observation of On-shift Activities

Based on real-time observational data across all 20 ED shifts, emergency physicians spent a median 34.1% of their shift on the computer [156.5 minutes (IQR 145.2–179.3)]. The longest interval of time away from the computer at any point during a shift was 20.0 minutes (range 8.0 to 20.0). Secondary to computer use, physicians spent a median of 26.9% of their shift with patients [115.2 minutes (IQR 102.0–154.1)] Among all 20 physicians, the longest time continuously spent at a patient’s bedside for any physician was 21m 47s (range 7.4 to 21.8 minutes).

Additionally, ED physicians spent a median of 15.9% of their shift [73.3 minutes (IQR45.6–96.4)] verbally communicating with staff. Physicians spent a median 9.5% of their total shift using the phone to talk to other healthcare workers [41.4 minutes (IQR 33.4– 59.2)], with a median duration of 1m 5s per phone task. Miscellaneous clinical tasks consisted of a median 5.5% [24.2 minutes (IQR 18.4–30.1)] of their shift. Clinical procedures included 0.7% EKG review [median 3.8 minutes (IQR 1.4–5.0)] and 0.4% other procedures [median 1.7 minutes (IQR 0.6–6.4)]. Time spent on EKG review is described in more detail in a related manuscript that focused on interruptions [29]. Across all physicians, personal time taken while working constituted only 3.9% of their total shift workflow [median 16.8 minutes (IQR 9.9–25.4)] and included time spent eating, using the restroom, and personal smartphone use.

Visualizations of physician workflow illustrate the substantial allocation of time spent on the computer compared to other clinical priorities as well as variations in workflow between and throughout ED shifts. *See Figures 3–5*. for visualizations of physicians’ time by activity (*Figure 3*), by individual shift (*Figure 4*), and linearly across each shift (*Figure 5*).

**Figure 3.**
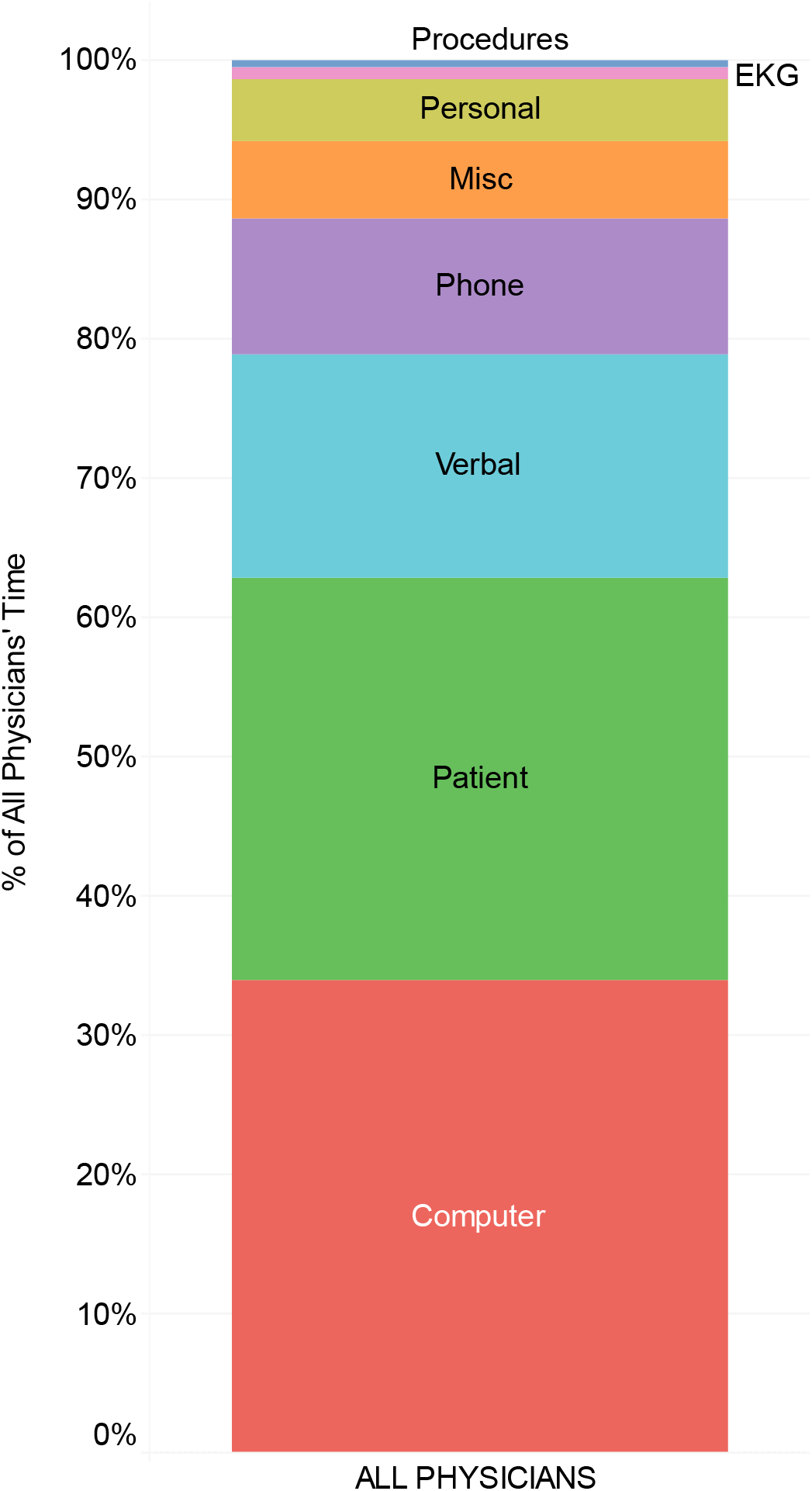
Allocation of Physician Time in the ED (Percent of Total Shift Across 20 Observed Physicians) Visualization illustrates cumulative allocation of physicians’ time on shift by activity category and across all observed physicians.

**Figure 4.**
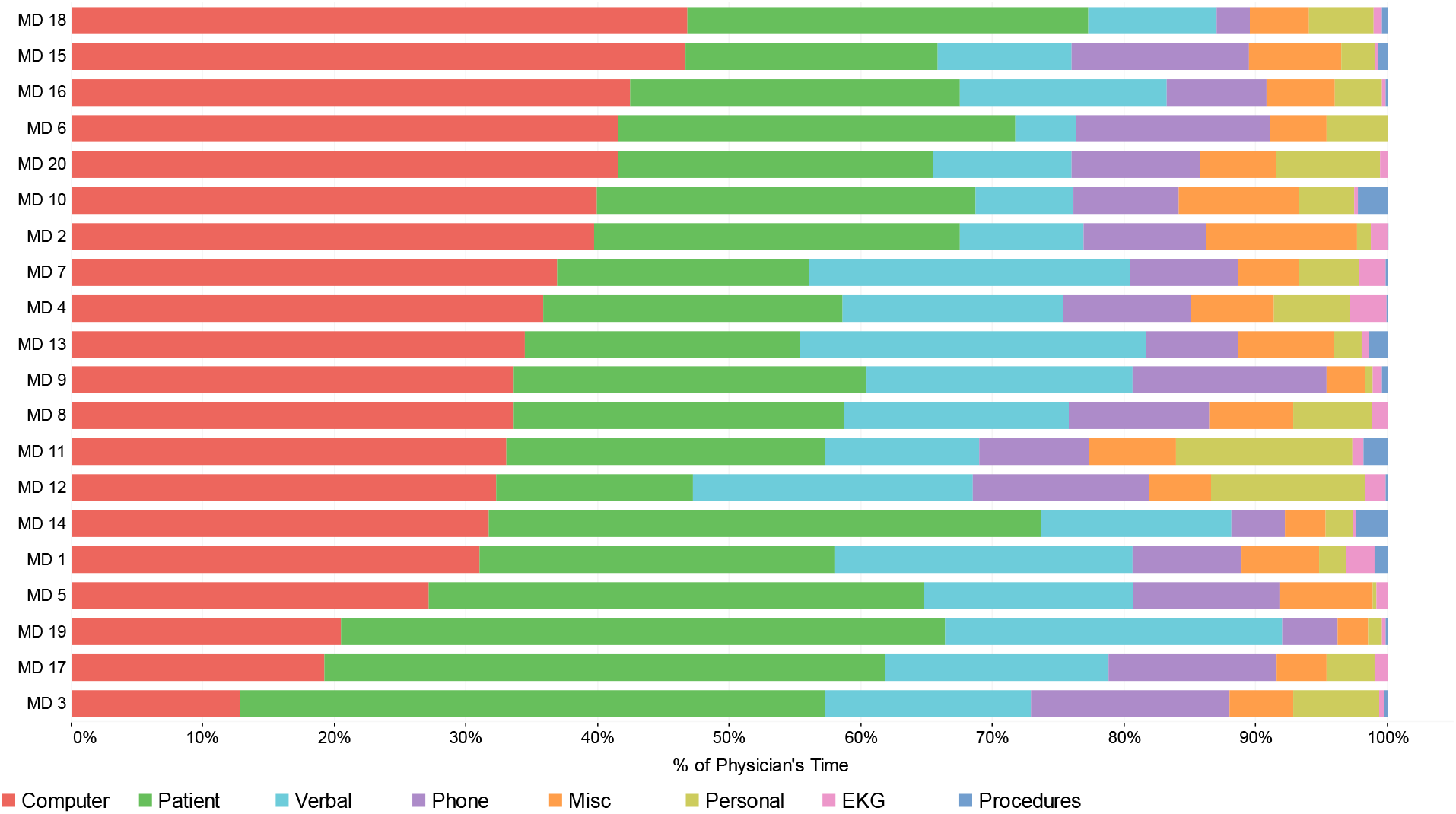
Allocation of Physician Time in the ED (By Physician) Visualization illustrates individual allocation of physicians’ time on shift by activity category and observed physician. Individual bars are sorted in descending order by percentage of Computer use.

**Figure 5.**
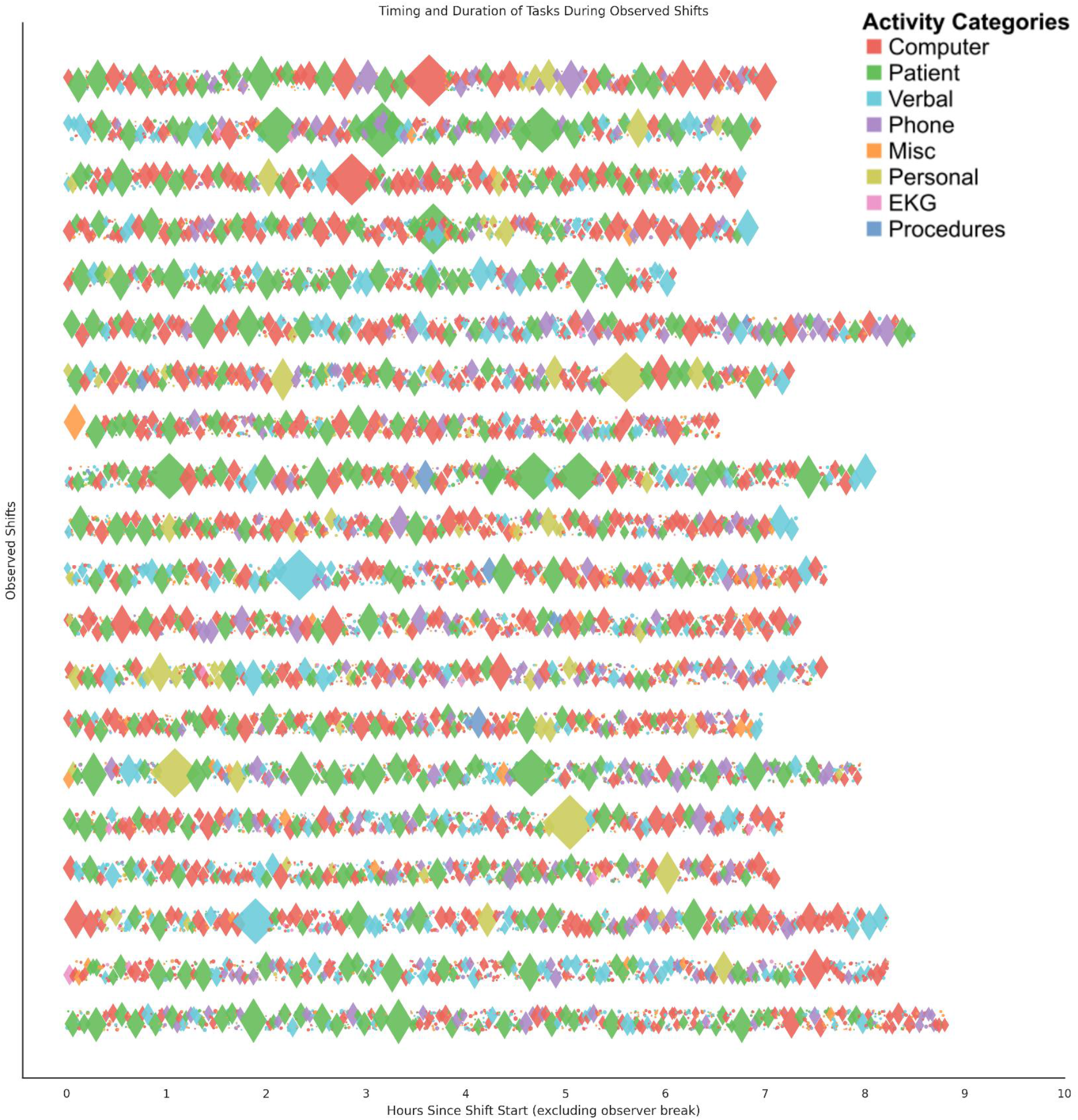
Timelines of Physician Workflow in the ED (By Physician) Visualization illustrates the sequence of activities during observed shifts from start to finish. The timeline of each shift flows horizontally across time with diamond-shaped markers scaled in size to represent the duration of each activity, which have been spread slightly with vertical ‘jitter’ for clarity. *\*Shifts (n=20) are sorted so the topmost has the fewest total number of tasks with observer’s 45 minute break omitted*.†*Diamond sizes are proportional to duration*.

#### After-Shift Computer Use

Across 19 clinical shifts, we obtained data on 112,396 EHR log events (actions) [median 5732 (IQR 5213–6519)] with a median event rate of 9.9 events per minute. The total EHR log timespan recorded from the log data, including active and idle computer use, was approximately 189.1 hours across 19 included physicians. Physicians spent a median 1.3h [(IQR 0.5–2.6); range 0.0h to 5.3h] for pajama time after the end of their scheduled shift.

## Discussion

In this pilot study of emergency physician workflow in a high-volume, metropolitan, Level 1 trauma center, our results reveal that physicians spent more time using computers than engaging in direct patient care — generally spending more than a third of their shift on the computer or approximately half of every scheduled hour when accounting for pajama time after their shift.

These findings add to a growing body of evidence that time spent on direct patient care is limited by competing workflow demands (especially burden of computer use) associated with emergency department care delivery [30-34].

This study combines data from both in-person time-motion observations and EHR event log data to demonstrate that the burden of computer use is not limited to time on-shift but also commonly extends hours beyond the end of clinical shifts [17, 19, 23]. While EHRs facilitate improved patient care by providing physicians access to patients’ prior medical histories and test results, for example, they also complicate the clinical work environment and may lead to unintended consequences such as limiting physician time at patients’ bedsides [33, 35].

Emergency physicians are required to adhere to documentation requirements, including written medical histories, physical examinations, interpretations of diagnostic test results, and medical decision-making notes [36]. While some documentation requirements have recently been revised to make them less burdensome, emergency physicians are still required to document clinical risk and complexity for billing purposes, which detracts from time available for direct patient care [37, 38].

Although current EHR systems facilitate access to essential clinical information, their user interface and design limitations may place an additional cognitive burden on emergency physicians [39-41]. Overly complicated interfaces and unintuitive action sequences in EHRs may frustrate physicians and place patients at risk for medical errors [42-44]. Pragmatic user-centered design principles should be incorporated as EHR systems are designed and tested, especially for those employed in ED care settings where time-critical actions are commonplace [33].

The development of more user-friendly interfaces that support seamless compliance and mitigate risk could significantly alleviate existing administrative pressures and allow physicians to focus more on patient-centered care [43, 45]. Developing these systems requires additional research to understand how clinicians’ varied current and future approaches to EHR use may impact the quality of care, for example by assessing whether more “staccato” (frequently interrupted) or, conversely, “legato” (fewer, longer periods of use) interactions are associated with increased efficiency or errors.

There is an urgent need for improved EHR design, documentation processes, and other human factors-centered solutions that enhance efficiency and improve quality and safety for patients [33, 46, 47]. Investing in “elbow-to-elbow” EHR training for emergency physicians may improve understanding of EHR functionalities, thereby enhancing their efficiency and reducing the associated burden for current and future clinicians [48-50].

Another emerging solution is the implementation of “ambient artificial intelligence scribes,” which use natural language processing to transform real-time audio-recordings of clinical encounters into a clinical note [51]. This technology is more promising than ever before due to rapid advancements in artificial intelligence (AI) capabilities over the last several years [33]. However, careful evaluation of such products must be undertaken to understand the strengths and limitations of the technology and identify unintended consequences for physicians (i.e., unexpected workflow challenges) as well as patients [52]. A recent article from a team at Kaiser Permanente expressed early enthusiasm for the use of ambient AI scribes, but they did note that the tool was not well-integrated into their EHR at the time of testing, which led to technical difficulties and frustrations due to wasted time [51]. Our study lays the groundwork for testing of tools such as AI scribes at our own site, since we now have baseline time-motion data that can be compared to future time-motion data collected during future tests of change utilizing innovative EHR and documentation tools.

### Limitations

Our study has several limitations. First, this was a single-site study, which may limit the generalizability of our results. Second, we recruited a convenience sample of attending physicians to participate in the study. As such, it is possible that physicians who opted to participate have systematic differences in the way they spend their time compared to those who did not participate. Third, because our observer was instructed not to enter patients’ rooms due to privacy concerns, we assumed that time in rooms was entirely spent on direct patient care. Thus, it is possible that we overestimated the amount of direct patient care and underestimated the amount of time spent on other activities such as computer use and phone calls that could have occurred within patient rooms. Fourth, to incorporate EHR event log data into our analyses, we selected inactivity period cutoffs times based on qualitative comparisons of EHR event log data and in-person observation data that nonetheless may systematically over- or underestimate EHR time; however, similar inactivity thresholds have been used for EHR event data in existing literature [15, 16]. Lastly, some physicians may have been using pajama time to catch up on documentation from prior shifts, which could have led to overestimation of total computer time. As we could not directly observe physicians after they left the ED, the use of EHR event logs with validated activity thresholds were found to be the most suitable approach for estimating post-shift EHR use that would have otherwise gone unmeasured [15, 16].

## Conclusions

This time-motion study found that emergency physicians allocate more than one-third of their time to computer use during their ED shifts. We analyzed EHR event logs to estimate that physicians also spend an additional 1–2 hours using the computer after their scheduled shift. In total, emergency physicians spend approximately half of every scheduled shift hour on computer-based work. Future research should investigate strategies to minimize low-yield time at the computer and increase time spent providing direct patient care.

## Data Availability

The datasets collected and analyzed in this study are not publicly available for protection of participants' privacy as they include potentially identifiable metadata and are governed by institutional data sharing policies. Deidentified data are available from the corresponding author upon reasonable request.

## Acknowledgements

This research was supported by internal funding from Cedars-Sinai Medical Center. The funding source did not play any role in study design, data collection, analysis, interpretation of results, writing, or the decision to submit this paper for publication.

## Author Contributions

CTB conceptualized the study. AJH conducted observations, oversaw data collection, and performed analyses. AJH, CTB, and KLI drafted the manuscript. All authors participated in writing and revision. AJH and CTB accept responsibility for the integrity of the work, maintained full access to the data, and initiated submission for publication.

